# Epidemiology of adverse events attributed to airway management in pediatric anesthesia: A protocol for a Japan Pediatric Difficult Airway in Anesthesia study (J-PEDIA)

**DOI:** 10.1101/2022.05.26.22275644

**Authors:** Taiki Kojima, Yusuke Yamauchi, Fumio Watanabe, Shogo Ichiyanagi, Yasuma Kobayashi, Yu Kaiho, Shugo Kasuya, Kevin Y. Urayama, Norifumi Kuratani, Yasuyuki Suzuki, J-PEDIA study collaborators

**Affiliations:** Department of Anesthesiology, Aichi Children’s Health and Medical Center, Aichi, Japan; Division of Comprehensive Pediatric Medicine, Nagoya University Graduate School of Medicine, Aichi, Japan; Children’s Heart Center, Saitama Children’s Medical Center, Saitama, Japan; Department of Anesthesiology, Tohoku University Hospital, Sendai, Japan; Department of Critical Care and Anesthesiology, National Center for Child Health and Development, Tokyo, Japan; Graduate School of Public Health, St. Luke’s International University, Tokyo, Japan; Department of Anesthesiology, Saitama Children’s Medical Center, Saitama, Japan

## Abstract

**Introduction:** Failure to secure an airway during general anesthesia is a major cause of adverse events (AEs) in children. The safety of peddiatric anesthesia may be improved by identifying the incidence of AEs and their attributed risk factors. However, previous large cohort studies have not been appropriately designed to explore such aspects. Thus, this study aims to deternine the incidence of AEs and the risk factors attributed to airway management under general anesthesia in children.

**Methods and design:** This prospective, multi-center, registry-based, observational study will be conducted in four tertiary care hospitals in Japan from June 2022 to May 2025. Children younger than 18 years of age undergoing surgical and/or diagnostic test procedures under general anesthesia or sedation by anesthesiologists will be enrolled in this study. Data on patient characteristics, discipline of anesthesia providers, and methodology of airway management will be collected through a standardized verification system under monitoring by institutional research leaders among the recruited institutions to minimize the loss of collected data. The primary outcome and exposure are AEs and presence of difficult airway features with potential confounders, which are related to the failure to secure the airway, and the variability in the anesthesia providers’ levels, adjusted using hierarchical multivariable regression models with mixed effects.

**Discussion:** This study will determine the incidence of AEs and the risk factors related to airway management under general anesthesia in children.

**Trial registration:** This study is registered as a prospective observational study in the University hospital Medical Information Network (UMIN) (registration number UMIN000047351, April 1, 2022).

## Introduction

The incidence of perioperative life-threatening adverse events (AEs) in children is higher than that in adults due to their unique physiological and anatomical characteristics [1]. These AEs are related to hypoxia in children commonly occur following failure to secure the airway since children are less tolerant to apnea [1-4]. Therefore, identifying the incidence of AEs and the risk factors in securing the airway is desired for safe and strategic airway management in pediatric anesthesia.

Epidemiological data on AEs during general anesthesia in children have been reported in previous large prospective cohort studies. The APRICOT study, which was the largest multi-center prospective study conducted in Europe, reported an incidence rate of 5.2% for severe perioperative events in children [5]. In addition, Subramanyam reported prediction models for perioperative respiratory adverse events based on preoperative patient comorbidities [6]. However, the APRICOT study was not designed to explore AEs specifically attributed to airway management during general anesthesia. Therefore, it did not include detailed information regarding the difficult airway features of patients, anesthesia methods, devices utilized for securing the airway, and disciplines of anesthesia providers. The PeDI registry study, a multi-center cohort study conducted in children’s hospitals in the US, reported that the incidence rate of intraoperative AEs was high in children with difficult airway features [2]. However, approximately 80% of the study cohort were patients with difficult airway features; therefore, the results did not represent the entire pediatric population. In addition, these major studies did not adjust for the variability in the techniques and skill level of anesthesia providers (e.g., each anesthesia provider could have a specific methodology for securing the airway and anesthesia). Therefore, the incidence of AEs and the related risk factors in securing the airway under general anesthesia in children remain unclear. In addition, to our knowledge, there is a lack of well-designed pediatric prospective studies using registry-based, real-world data in the Asian region regarding the incidence of AEs and the risk factors attributed to airway management.

This study aims to investigate the incidence of AEs attributed to airway management during general anesthesia in children and to identify their risk factors.

## Methods

### Study design and setting

This is a prospective, registry-based, real-world, multicenter, observational study that will initiate data registration from June 1, 2022, to May 31, 2025, in five tertiary care hospitals in Japan: Aichi Children’s Health and Medical Center (Aichi), National Center for Child Health and Development (Tokyo), Saitama Prefectural Children’s Medical Center (Saitama), Tohoku University Hospital (Sendai), and Hokkaido University Hospital (Sapporo).

### Target population

#### Inclusion criteria

We will enroll cases that meet all of the following criteria.

1. Children younger than 18 years of age who receive advanced airway management under general anesthesia or sedation for surgical and/or test procedures conducted by anesthesiologists or anesthesia providers under the supervision of anesthesiologists.
2. General anesthesia or sedation is performed in operating suites, catheterization laboratory rooms, rooms for radiological imaging and procedures (e.g., computed tomography, magnetic resonance imaging, radiation therapy), or general ward.

#### Exclusion criteria

We will exclude cases that meet at least one of the following criteria.

– Cases outside operating suites where anesthesiologists are consulted for airway management.
– Cases where airway management is performed in the emergency department and intensive care unit.
– Duplicate cases during the study period.
– Patients or their families who refuse to participate in the study for any reason.

### Data collection

This registry-based observational study will prospectively collect data regarding the characteristics of patients and surgery, disciplines of anesthesia providers, practice of airway management, occurrence of AEs, and treatments for AEs.

#### Data quality control

Data will be initially collected by anesthesia providers for each attempt to secure the airway with paper-based data collection forms. The collected initial paper-form-based data will be verified by each site-specific research leader through a standardized process among the four recruited institutions. Site-specific research leaders will confirm that the capture rate is ≥95% of cases at each institution. To standardize the definitions of the terms for data collection, they are predetermined and described in the operational research manual created by the primary investigator (TK). The research operational committee will be convened regularly to confirm the definitions of the terms used in the data collection process. Site-specific research leaders instructed the anesthesia providers to increase the accuracy of the collected data at each recruited institution, aiming to minimize misclassification bias (Figure 1).

#### Data storage and process

The initially collected p≥aper-form-based data will include information that can identify the study participants (i.e., medical record number and name initials). Thus, only anonymous information will be registered in the Research Electronic Data Capture (REDCap^®^, Tokyo, Japan) [7] system hosted by the National Center for Child Health and Development. Site-specific research leaders will utilize a standardized participant’s correspondence file, which will include the participant’s initials, medical record number, and REDCap registration number for a potential review (Figure 1).

**Figure 1.**
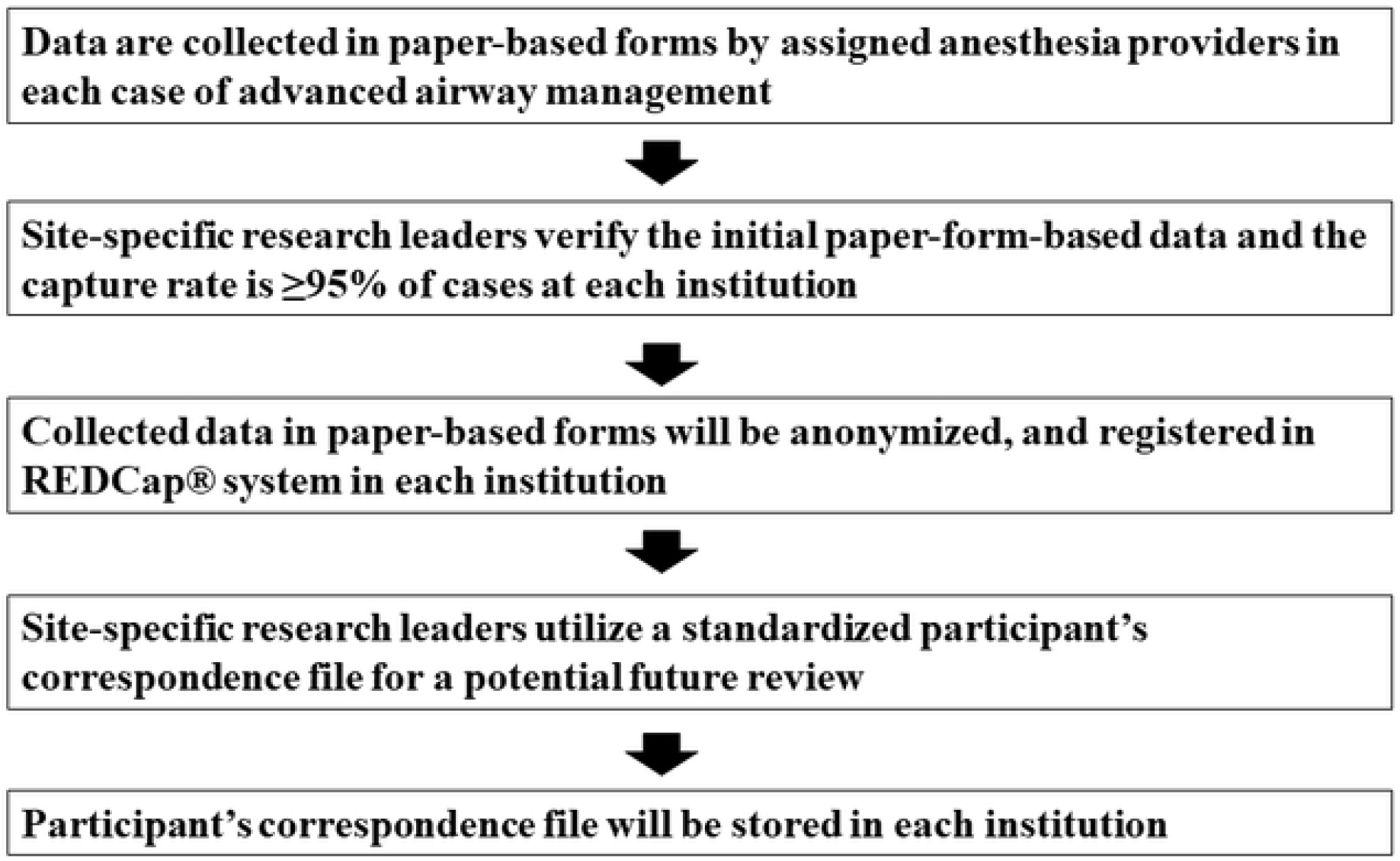
Flow of collection, quality control, and storage of data

#### Endpoints

The primary and secondary endpoints are any AEs attributed to securing the airway and decreased values of peripheral capillary oxygen saturation (SpO_2_) from the time of initiation to completion of securing the airway. The criteria for AEs were predetermined [i.e., cardiac arrest (status of dead or alive at 24 hours after the critical event happened), laryngospasm, upper airway obstruction, severe cough, bronchial intubation, esophageal intubation, vomiting, hypotension, hypertension, tooth injury, pneumothorax, mediastinal emphysema, bronchospasm, atelectasis, pulmonary edema, stridor, airway trauma, arrhythmia, agitation, dislodgement of airway securing devices] based on the National Emergency Airway Registry for children (NEAR4KIDS), which is a national registry database to evaluate AEs during emergency tracheal intubation in pediatric intensive care units mostly located in North America [8]. The AEs have been classified as severe and non-severe before data collection. Severe AEs include cardiac arrest, esophageal intubation with a ≥ 5% decrease in SpO_2_, vomiting with aspiration, hypotension requiring treatments, laryngospasm, pneumothorax, mediastinal emphysema, bronchospasm, pulmonary edema, and direct airway trauma. Non-severe AEs include bronchial intubation, esophageal intubation with a < 5% decrease in SpO_2_, vomiting without aspiration, hypertension requiring treatments, upper airway obstruction, severe cough, tooth injury, atelectasis, stridor, arrhythmia, and dislodgement of airway securing devices.

#### Exposures and potential confounders

Exposure is defined as the presence of any of the difficult airway features (i.e., preoperative recognition of difficult airway by anesthesiologists, history of difficult airway, restricted cervical flexion, restricted mouth opening, narrow thyromental distance, upper airway obstruction, or anatomical barrier to visualize glottic opening, midfacial hypoplasia, large tongue, micrognathia, and large head circumference) that were predetermined based on previous studies [9]. Potential confounders were selected before initiation of data collection based on a review of previous literature and clinical experience of experienced anesthesiologists among the research team members. The selected potential confounders were age, sex, prematurity, obesity (weight-for-age ≥95 percentile), preoperative comorbidities, families smoking status, provider discipline, location, institution, fiscal year, Cormack-Lehan grade, method of anesthesia induction, status of nothing per os and a full stomach, type of device for securing the airway, and application of external laryngeal manipulation or cricoid pressure [9-14].

### Statistical considerations

#### Sample size estimation

The reported incidence rate of critical events occurring during general anesthesia induction in children was 5.2%, including an incidence of 1.9% for cardiovascular instability in the general pediatric population who received general anesthesia [5]. Therefore, we estimated the sample size with the assumption of the lowest incidence of critical adverse events during general anesthesia in children as 2.0%. The estimated sample size was approximately 16,000 assuming a 99% probability of obtaining a 95% Wilson CI with ± 0.3% of the half-width for the incidence of critical adverse events.

#### Statistical analysis

For summary statistics, categorical variables will be described as numbers and percentages, whereas normally and non-normally distributed continuous variables will be described as means and standard deviations or medians and interquartile ranges. Univariable and multivariable analyses will be performed to evaluate risk factors associated with the primary outcome. Baseline differences between subjects with and without difficult airway features will be evaluated using the Student’s t-test for normally distributed continuous variables, Wilcoxon rank-sum test for continuous variables with skewed distributions, and chi-square test or Fisher’s exact test for categorical variables. Multilevel logistic regression models with mixed effects will be used to evaluate the association between the occurrence of adverse events while securing the airway and the presence of difficult airway features [15]. Hierarchical relative risk regression models will be developed using robust error variances or hierarchical logistic regression models for multivariable analysis. Potential confounders for patient, anesthesia, provider characteristics, and institutions will be selected based on clinical relevance and adjusted for fixed effects. The variance contributed by the clustering effects based on the different clinical practices among anesthesia providers will be adjusted (each subject as level 1, each anesthesia provider as level 2) as random effects [15]. Cases with missing values will be removed from the analysis. Data will be analyzed using STATA 17.0 (StataCorp, College Station, TX, USA), with a two-sided *p*-value of <0.05, serving as the criterion for assessing the null hypothesis for each analysis.

### Ethics statement

This study will be conducted in accordance with the declaration of Helsinki. The study protocol was reviewed and approved by the institutional review board at Aichi Children’s Health and Medical Center (approval number 2021051, September 29, 2021) and local ethical approval will be obtained from each recruited institution before initiating data collection. The institutional review board considered that written consent could be waived by performing the opt-out procedure in the current clinical study.

### Dissemination

The results of this study will be reported in a peer-reviewed journal and a relevant cademic conference.

### Harms

No harmful events will occur during data collection because of the nature of an observational study without interventions.

## Discussion

This is a prospective, registry-based, multicenter, observational study that aims to describe real-world data on the incidence of AEs and the risk factors attributed to securing the airway in children under general anesthesia. Investigating real-world data regarding patient characteristics (including the prevalence of difficult airway features) and characteristics of pediatric anesthesia practice can provide anesthesia providers with useful information to improve the safety and effectiveness of airway management.

This study has several important limitations owing to its observational nature of the study. First, unmeasured confounders could remain unadjusted owing to the observational nature of the study. Second, there might be a selection bias due to the sampling method. Third, reporting bias (i.e., misclassification bias) can occur due to incorrect reports during data collection. To address these potential limitations, we carefully designed this study to collect detailed information regarding the exposure (presence of difficult airway features), potential confounders, and institutions based on previous literature and meetings with experienced board-certified anesthesiologists during the development of the study protocol. In addition, we structured a standardized system to confirm the data collected by local research leaders among the recruited institutions. This standard data confirmation system has been used in NEAR4KIDS, which achieved a data capture rate of ≥95% [11]. Our standardized data collection system allows our study to maintain the quality of the collected data and minimize potential information biases (e.g., misclassification). In addition, the REDCap^®^ data registration system does not allow data registration to proceed if there are missing data. We will use this system for data registration to minimize missing data for future analysis.

Previous registry-based prospective studies (i.e., APRICOT study, PeDI registry study) were performed mainly in Europe and North America [2,5]. The evidence regarding the incidence of AEs related to securing the airway is insufficient in Asian settings. The incidence and risks of AEs attributed to airway management in the Japanese pediatric population might be different from those in Europe and North America owing to the differences in the characteristics of the patient population and anesthesia practices. This prospective, registry-based, real-world observational study in Japan can be a stepping stone for further research to identify the specific risk factors for AEs attributed to failure to secure the airway in the Asian region.

## Data Availability

No datasets were generated or analysed during the current study. All relevant data from this study will be made available upon study completion.

## Abbreviations

AE: Adverse event
NEAR4KIDS: National Emergency Airway Registry for children

## Ethics statements

### Patient consent for publication

Not required.

## Acknowledgments

None.

## Author Contributions

**Conceptualization:** Taiki Kojima, Yusuke Yamauchi, Fumio Watanabe, Shogo Ichiyanagi, Yasuma Kobayashi, Yu Kaiho, Shugo Kasuya, Norifumi Kuratani and Yasuyuki Suzuki

**Data curation:** Taiki Kojima, Yusuke Yamauchi

**Funding acquisition:** Taiki Kojima [Grants-in-Aid for Scientific Research (Kakenhi Grant) #22K09085, April 1, 2022].

**Methodology:** Taiki Kojima, Yusuke Yamauchi, Fumio Watanabe, Shogo Ichiyanagi

**Writing - original draft:** Taiki Kojima

**Writing – review & editing:** Taiki Kojima, Yusuke Yamauchi, Fumio Watanabe, Shogo Ichiyanagi, Yasuma Kobayashi, Yu Kaiho, Shugo Kasuya, Kevin Y. Urayama, Norifumi Kuratani, Yasuyuki Suzuki

## References

1. Else SDN, Kovatsis PG. A narrative review of oxygenation During pediatric intubation and airway procedures. Anesth Analg. 2020;130: 831–840. doi: 10.1213/ANE.0000000000004403.

2. Fiadjoe JE, Nishisaki A, Jagannathan N, Hunyady AI, Greenberg RS, Reynolds PI, et al. Airway management complications in children with difficult tracheal intubation from the Pediatric Difficult Intubation (PeDI) registry: A prospective cohort analysis. Lancet Respir Med. 2016;4: 37–48. doi: 10.1016/S2213-2600(15)00508-1.

3. Schibler A, Hall GL, Businger F, Reinmann B, Wildhaber JH, Cernelc M, et al. Measurement of lung volume and ventilation distribution with an ultrasonic flow meter in healthy infants. Eur Respir J. 2002;20: 912–918. doi: 10.1183/09031936.02.00226002.

4. Humphreys S, Pham TM, Stocker C, Schibler A. The effect of induction of anesthesia and intubation on end-expiratory lung level and regional ventilation distribution in cardiac children. Paediatr Anaesth. 2011;21: 887–893. doi: 10.1111/j.1460-9592.2011.03547.x.

5. Habre W, Disma N, Virag K, Becke K, Hansen TG, Jöhr M, et al. Incidence of severe critical events in paediatric anaesthesia (apricot): A prospective multicentre observational study in 261 hospitals in Europe. Lancet Respir Med. 2017;5: 412–425. doi: 10.1016/S2213-2600(17)30116-9.

6. Subramanyam R, Yeramaneni S, Hossain MM, Anneken AM, Varughese AM. Perioperative respiratory adverse events in pediatric ambulatory anesthesia: Development and validation of a risk prediction tool. Anesth Analg. 2016;122: 1578–1585. doi: 10.1213/ANE.0000000000001216.

7. Harris PA, Taylor R, Thielke R, Payne J, Gonzalez N, Conde JG. Research Electronic Data Capture (REDCap)--A metadata-driven methodology and workflow process for providing translational research informatics support. J Biomed Inform. 2009;42: 377–381. doi: 10.1016/j.jbi.2008.08.010.

8. Branca A, Tellez D, Berkenbosch J, Rehder KJ, Giuliano JS, Gradidge E, et al. The New Trainee Effect in Tracheal Intubation Procedural Safety Across PICUs in North America: A Report From National Emergency Airway Registry for Children. Pediatr Crit Care Med. 2020;21: 1042–1050. doi: 10.1097/PCC.0000000000002480.

9. Heinrich S, Birkholz T, Ihmsen H, Irouschek A, Ackermann A, Schmidt J. Incidence and predictors of difficult laryngoscopy in 11,219 pediatric anesthesia procedures. Paediatr Anaesth. 2012;22: 729–736. doi: 10.1111/j.1460-9592.2012.03813.x.

10. Valois-Gómez T, Oofuvong M, Auer G, Coffin D, Loetwiriyakul W, Correa JA. Incidence of difficult bag-mask ventilation in children: A prospective observational study. Paediatr Anaesth. 2013;23: 920–926. doi: 10.1111/pan.12144.

11. Kojima T, Laverriere EK, Owen EB, Harwayne-Gidansky I, Shenoi AN, Napolitano N, et al. Clinical impact of external laryngeal manipulation during laryngoscopy on tracheal intubation success in critically ill children. Pediatr Crit Care Med. 2018;19: 106–114. doi: 10.1097/PCC.0000000000001373.

12. Mirghassemi A, Soltani AE, Abtahi M. Evaluation of laryngoscopic views and related influencing factors in a pediatric population. Paediatr Anaesth. 2011;21: 663–667. doi: 10.1111/j.1460-9592.2011.03555.x.

13. Akpek EA, Mutlu H, Kayhan Z. Difficult intubation in pediatric cardiac anesthesia. J Cardiothorac Vasc Anesth. 2004;18: 610–612. doi: 10.1053/j.jvca.2004.07.003.

14. Graciano AL, Tamburro R, Thompson AE, Fiadjoe J, Nadkarni VM, Nishisaki A. Incidence and associated factors of difficult tracheal intubations in pediatric ICUs: A report from National Emergency Airway Registry for Children: NEAR4KIDS. Intensive Care Med. 2014;40: 1659–1669. doi: 10.1007/s00134-014-3407-4.

15. Austin PC. A tutorial on multilevel survival analysis: Methods, models and applications. Int Stat Rev. 2017;85: 185–203. doi: 10.1111/insr.12214.

